# Retinal optical coherence tomography angiography imaging in population studies

**DOI:** 10.1101/2025.01.22.25320610

**Authors:** Amir H Kashani, Tos TJM Berendschot, Sophie Bonnin, Lucia Sobrin, Danilo Andrade De Jesus, Luisa Sanchez Brea, Ana Collazo Martinez, Rajiv Raman, Victor A de Vries, Haifan Huang, E3 consortium, Maastricht Study, Framingham Heart Study, Rotterdam Study, Rhineland Study, SIGNATR Study, Tom MacGillivray, Wishal D Ramdas, Frank CT van der Heide, Carroll A Webers, Freekje van Asten, Alexandra Beiser, Sudha Seshadri, Caroline Klaver, Monique Breteler, Robert Finger, Rehana Kahn, Renee Liu, Gayatri Susarla, Azzi N Rizza, Ashley Li, Samuel Han, Weilin Chan, Ines Lains, Janine Yang, Kim Brustoski, Vikas Khetan, Sudha Iyengar, Penelope Benchek, Sinnakaruppan Mathavan, Ricky Chan

## Abstract

**Background:** Widespread use of retinal optical coherence tomography angiography (OCT-A) imaging in population-based studies requires methodological and analytical consensus to ensure reproducible and accurate results.

**Objective:** Development of a consensus framework for assessment of OCT-A image quality and working with OCT-A-derived variables in population studies.

**Methods:** Criteria for image quality were developed for fovea-centered, 3×3-mm OCT-A images acquired with three commercially available devices. Inter- and intragrader agreements for overall image quality, vessel density (VD) and foveal avascular zone (FAZ) area were evaluated among five graders (% agreement). Intergrader agreement was validated on 6×6-mm OCT-A images. Recommendations for grading and use of OCT-A images in epidemiological studies were developed.

**Results:** Mean intergrader agreements for overall “Unusable”, “Usable”, and “Excellent” 3×3-mm OCT-A image quality (52 images) were 82.3%, 70.8%, and 87.1%; for “Unusable” VD and FAZ area agreements were 80.0% and 81.9%, respectively. Mean intragrader agreements for overall “Unusable”, “Usable” and “Excellent” 3×3-mm OCT-A image quality (27 images) were 91.1%, 82.2%, and 81.9%; agreements for “Unusable” VD and FAZ area were 89.6%; and 92.6%, respectively. Mean intergrader agreements for assessment of overall “Unusable”, “Usable” and “Excellent” 6×6-mm OCT-A images (21 images) were 73.3 %, 57.1% and 83.8%, agreements for “Unusable” VD and FAZ area were 60.0 % and 84.8 %, respectively. Three analytic scenarios were developed to account for common types of bias related to suboptimal OCT-A image quality and ocular comorbidities.

**Conclusion:** These recommendations provide a framework for working with OCT-A imaging in population studies.

## Introduction

The advent of retinal optical coherence tomography angiography (OCT-A) enables a revolutionary opportunity for imaging the retinal microvasculature.^1, 2^ With OCT-A superficial and deep retinal capillary networks can be quantified *in vivo*, non-invasively, and with high accuracy (up to a semi-histological resolution).^1^ Common measures extracted from OCT-A images are capillary density (e.g. vessel skeleton density) and foveal avascular zone area size.^1^ Lower capillary density and larger foveal avascular zone area are considered to reflect worse capillary and systemic health.^1, 2^

There is an increasing interest in the use of OCT-A imaging in population-based studies. Indeed, the acquisition of OCT-A images has started in multiple large cohort studies, including the Maastricht Study (the Netherlands), Rotterdam Study (the Netherlands), Rhineland study (Germany), Framingham Heart study (USA), and South Indian GeNetics of DiAbeTic Retinopathy (SIGNATR) Study (India). Such studies allow the large-scale study of chronic diseases with a microvascular origin, including ocular diseases (such as glaucoma,^3^ diabetic retinopathy,^4^ and age-related macular degeneration^5^) and non-ocular diseases (such as neurodegenerative diseases,^6^Alzheimer’s disease,^7^ stroke,^8^ and heart failure^9^). The biological grounding for studying non-ocular diseases is that retinal capillary health (in part) reflects systemic capillary health.^9^

A standardized approach for working with OCT-A data in population studies is needed to promote use of optimal and similar methods across cohorts internationally.^4^ Such an approach should ideally consider bias which may arise due to suboptimal OCT-A image quality, e.g. information bias and selection bias.^4, 10^ For example, if less healthy individuals more often have poor image quality and are more likely to be excluded from analyses, an underestimation of the association may occur.^10^

No methodological framework for working with OCT-A images in population studies has yet been widely adopted. One study developed criteria for grading OCT-A image quality in neurological studies (OSCAR-MP criteria),^11^ but further development of criteria is required for use in population studies. Limitations of this previous study are that 1) no criteria specific for key OCT-A measures such as vessel density and foveal avascular zone area were developed; 2) criteria were only developed on one type of scanning protocol from one OCT-A imaging device; and 3) performance was only evaluated among patients with neurological diseases (and not in the general population).^11^

In view of the above, researchers from the European Eye Epidemiology consortium (The Maastricht Study, Rotterdam Study, Rhineland Study) initiated a shared effort to develop recommendations for working with OCT-A images in population-based studies. Invited were also researchers from ongoing collaborations, including researchers from the Framingham Heart Study, SIGNATR, and the Adolphe de Rothschild Foundation Hospital. Overarching aim was to develop a reliable and reproducible framework for assessment of OCT-A image quality and conducting analyses with OCT-A-derived variables in population-based studies.

## Methods

### Part 1: Design of quality criteria for grading fovea-centered OCT-A images

We aimed to develop a minimum set of image quality criteria that could simply and quickly be assessed and has a high reproducibility (i.e. high inter- and intragrader agreement), for use in population-based studies. We consider quality of the overall image and usability of an image to determine capillary density and foveal avascular zone area.

#### Study population and design

Prospectively collected data from the following four observational cohort studies were used: Maastricht Study (The Netherlands),^12^ Rotterdam Study (The Netherlands),^13^ Framingham Heart Study (USA),^14^ and SIGNATR (India).^15^ General characteristics of the study populations among which OCT-A imaging was conducted are shown in Table 1. Mean age ± standard deviation and % men are: 68 ± 8 years and 51% men in Maastricht Study (n=2,567 [collected by May 1st 2024]); 75 ± 7 years and 43% men in Rotterdam Study (n=2,136 [collected by July 17th 2024]); 75 ± 7 years and 41% men in Framingham Heart Study (n=962 [collected by 26^th^ of August 2024]); and 57 ± 10 years and 54% men in SIGNATR (n=348 [collected before July 1st 2024]). In addition, data collected among 5 healthy individuals (1-man, 4 women age range 30-62 years old) at Rhineland Study; and 4 healthy women (age range 28-39 years old) from Adolphe de Rothschild Foundation Hospital were used. For all cohort studies medical ethical approval was obtained. All participants provided informed consent. More details on cohort studies are presented in the Supplemental Material 1.

**Table 1.** Characteristics of the studies and research centers from which OCT-A images were used.

|  | Maastricht Study | Rotterdam Study | Framingham Heart Study | Rhineland study | SIGNATR | Adolphe de Rothschild Foundation Hospital |
| --- | --- | --- | --- | --- | --- | --- |
| **Study characteristics** |  |  |  |  |  |  |
| Country | The Netherlands | The Netherlands | USA | Germany | India | France |
| Design | Population study (oversampling of type 2 diabetes) | Population study | Population study | Population study | Type 2 diabetes study | Clinical patients from ophthalmology hospital department |
| Total sample with OCT-A images* | 2,567 | 2,136 | 962 | 3,472 | 348 | NA |
| Mean age (SD) | 68 (8) | 75 (7) | 75 (7) | 59 (13) | 57 (10) | NA |
| Men | 51% | 43% | 41% | 43% | 54% | NA |
| **OCT-A image characteristics** |  |  |  |  |  |  |
| Device brand | Heidelberg Engineering | Topcon | Zeiss | Heidelberg | Zeiss | Zeiss |
| Model | Spectralis | Triton | Angioplex | Spectralis | HD-OCTA | Angioplex |
| Protocol | 3x3-mm | 3x3-mm | 3x3-mm | 3x3-mm | 6x6-mm | 3x3-mm; 6x6-mm |
| Software | Heidelberg Eye Explorer 1.11.2.0 | IMAGEnet6 v1.04E | Cirrus Review 11.5.2.54532 | Heidelberg Eye Explorer 1.11.2.0 | Cirrus Review 11.5.2.54532 | Cirrus Review 11.5.2.54532 |
| Number of B-scans for 3x3-mm protocol | 512 | 320 | 245 | 512 | 245 | NA |
| Number of B-scans for 6x6-mm protocol | NA | NA | NA | NA | 350 | 350 |
| **Images used for image quality criteria development & validation** |  |  |  |  |  |  |
| Number of images used |  |  |  |  |  |  |
| -preliminary discussion | 3 | 3 | 3 | 3 | 3 | 3 |
| -criteria development | 10 | 10 | 10** | 10 | 10 | 10** |
| -Intergrader agreement*** | 12 | 10 | 10** | 10 | 10 | 10** |
| -Intragrader agreement*** | 9 | 4 | 3 | 6 | NA | NA |

\*Numbers available by up to August 2024; \*\*indicates the same images were used; \*\*\*For inter- and intragrader agreement the same images were used. Abbreviations: NA, not applicable.

#### OCT-A image acquisition

Four OCT-A devices were used to acquire fovea-centered images: Spectralis (Heidelberg Engineering, Heidelberg, Germany), used in Maastricht Study and Rhineland Study; Triton (Topcon, Tokyo, Japan), used in Rotterdam Study; Cirrus Model 5000 (HD-OCTA, Carl Zeiss Meditec, Inc., Dublin, CA, USA;) used in SIGNATR and at the Adolphe de Rothschild Foundation Hospital; and Cirrus Model 6000 (AngioPlex; Carl Zeiss Meditec, Inc., Dublin, CA, USA), used in the Framingham Heart Study. All these are common devices used, allowing to develop criteria that can be applied in multiple studies. Fovea-centered OCT-A scans with dimensions of 3×3-mm were available in all studies except for SIGNATR. 6×6- mm OCT-A images were available only in the SIGNATR cohort and at the Adolphe de Rothschild Foundation Hospital. We used full-slab OCT-A images (showing both superficial and deep capillary networks) for all assessments. Exemplary OCT-A images from all centers are shown in Figures S1 and S2 (Supplemental Material 1).

Number of B-scans acquired (software version) for 3×3-mm OCT-A images were: 512 (Heidelberg Eye Explorer 1.11.2.0) in Maastricht Study and Rhineland Study; 320 (IMAGEnet6 v1.04E) in Rotterdam Study; 245 (Cirrus Review 11.5.2.54532) in Framingham Heart Study and 245 (Cirrus Review 11.5.2.54532) at Adolphe de Rothschild Foundation Hospital. Number of B-scans acquired (software version) for 6×6-mm OCT-A images was: 350 (Cirrus Review 11.5.2.54532), both in SIGNATR (HD-OCT) and at Adolphe de Rothschild Foundation Hospital.

#### Multidisciplinary team

Recommendations were developed within a multicenter, multidisciplinary team consisting of ophthalmologists, optometrists, and researchers with backgrounds in clinical epidemiology, optical physics, and medical image data science. This approach enables consideration of key technical, biological, and epidemiological analyses-related factors. Ophthalmologists are: A.H.K. and L.S. (Framingham Heart Study), W.D.R. (Rotterdam Study), H.H. (Rhineland Study), and S.B. (Fondation Adolphe de Rothschild Foundation Hospital); clinical epidemiological researchers are: F.C.T.v.d.H. (Maastricht Study and Adolphe de Rothschild Foundation Hospital), V.A.V. (Rotterdam Study), L.S. (Framingham Heart Study), and A.C. (Framingham Heart Study) and; researcher in the field of optical physics is T.B. (Maastricht Study); and medical image data scientist are D.A.J. and L.S.B. (Rotterdam Study). Contributions of all other authors are listed in the author contributions section.

#### Development of image quality criteria

Criteria were initially developed using 3×3-mm OCT-A scans because it was the most commonly used type of protocol across cohorts and is the highest resolution scan protocol on all devices. We used a three-level grading system (“Excellent,” “Usable,” “Unusable”) to specify quality of the overall image; and a two-level system (“Usable”, “Unusable”) to specifically determine the usability of vessel density and foveal avascular zone area for quantitative measures. These variables were selected as they are the most commonly used OCT-A-derived measures.^1^ A separate classification for foveal avascular zone area was used because in some cases the quality of the scan in that region was sufficient for quantification, but the majority of the image was not suitable for quantification or vice-versa.

Development of quality criteria took place via a three-stage process during bimonthly teleconferences involving researchers from all centers over a period of six months. In the <u>first</u> <u>stage,</u> a preliminary categorization of images was made, classifying into “Excellent,” “Usable,” and “Unusable”. For this purpose, individual researchers from each center independently provided three images from their data that were thought to be representative of these qualities. In the <u>second stage,</u> the group identified images that could serve as exemplary images for each category of image quality by group discussion and consensus; and we formulated a definition for each category of image quality. Fifty images obtained from all centers were used for the second stage, with each center providing 10 randomly selected images. In the <u>third stage,</u> we evaluated the reliability of the definitions from the second stage by quantifying inter- and intragrader agreement. Inter- and intragrader agreement among and within five graders was determined using 52 and 27 images, respectively. The same images from Adolphe de Rothschild Foundation Hospital and Framingham Study were used for both the second and third stage (20 images); and different (randomly selected) images from the Maastricht Study, Rhineland Study, and Rotterdam Study were used in the second and third stages (30 for the second stage and 32 for the third stage). OCT-A images were presented in a random order (i.e. not per center or imaging device). For assessment of intragrader agreement, 12 of the 27 images that were presented a second time were randomly rotated by 90 or 180 degrees to reduce the chance graders would recognize the image. Agreement was quantified as percentage agreement.^10^ For three-level classification “Excellent”, “Usable”, “Unusable”, one category was compared with both other categories combined, e.g. “Unusable” versus “Usable” or “Excellent”.

#### Validation of image quality criteria on a 6×6-mm OCT-A image

Intergrader agreement among the same five graders was assessed using 21 randomly selected images from SIGNATR and Adolphe de Rothschild Foundation Hospital. Intergrader agreement was quantified as percentage agreement.^10^

#### Additional analyses

For comprehensiveness and comparison with literature,^11^ we also calculated agreement using Cohen’s Kappa.^16, 17^ We did not use Cohen’s Kappa as the main measure of agreement for the following reason. The calculation of percentage agreement depends on grader agreement only, but calculation of Cohen’s kappa depends on both grader agreement and distribution of samples across categories. As the use of small sample sizes increases the risk of unequal distributions across subgroups, Cohen’s kappa may provide a less robust measure of intergrader agreement than percentage agreement. ^16, 17^

#### Part 2: Framework for data analyses

We aimed to develop a framework for working with OCT-A images in population-based studies. Requirements were that this framework should 1) enable insight into how suboptimal OCT-A image quality may affect effect size estimations (whilst considering both information and selection bias); and 2) specify how to deal with common ocular comorbidities.^10^

## Results

### Part 1: Design of quality criteria for grading of fovea-centered OCT-A images

#### Criteria development

After preliminary discussions (stage one), exemplary images were selected and definitions for grading image quality were developed (stage two). Figure 1 shows exemplary OCT-A images for each category of image quality; and panel 1 shows image quality descriptions.

**Panel 1**

**“Excellent” image quality**

Excellent discrimination of the “capillary vasculature” throughout the entirety of the OCT-A scan and clear demarcation of the foveal avascular zone edges. The fovea is positioned in the centre of the image (i.e. the geometric center of the fovea is within 1 foveal diameter of the image field-of-view center).

**“Usable” image quality for quantification of vessel density**

Discrimination of “capillary vasculature” may be less distinct than “Excellent” but still of sufficient quality for acquisition of reliable data in the majoritya of the image. Some image artefacts can be accepted (e.g. non-continuousb arterioles/venules). The fovea is positioned in the centre of the image as above.

**“Usable” image quality for quantification of foveal avascular zone area**

Discrimination of foveal avascular zone edges may be less distinct than “Excellent” but still of suitable quality to assess foveal avascular zone area or perimeter (there should be no [motion] artefacts in the ring). The inner capillary ring is manually traceable with minimal subjective interpolation needed.

**“Unusable” image quality**

Discrimination of the “capillary vasculature” is not distinct on the largest parta of the image and foveal avascular zone edges are not clear (demarcation of the edges may not be clear due to motion artefacts, projection artefacts, or any other reason).

Footnotes:

^a^Largest part of the image refers to a subjective assessment that at least 80% of the image, assessed upon visual inspection, is of “Usable” quality. This cut-off was chosen as some researchers expressed their wish for a clear cut-off, but acknowledge that purely objective quantification may be too labor intensive for practical implementation in large studies.

^b^Non-continuous indicates “large vessel that is broken by full width” (an example is provided in Figure 2).

**Figure 1.**
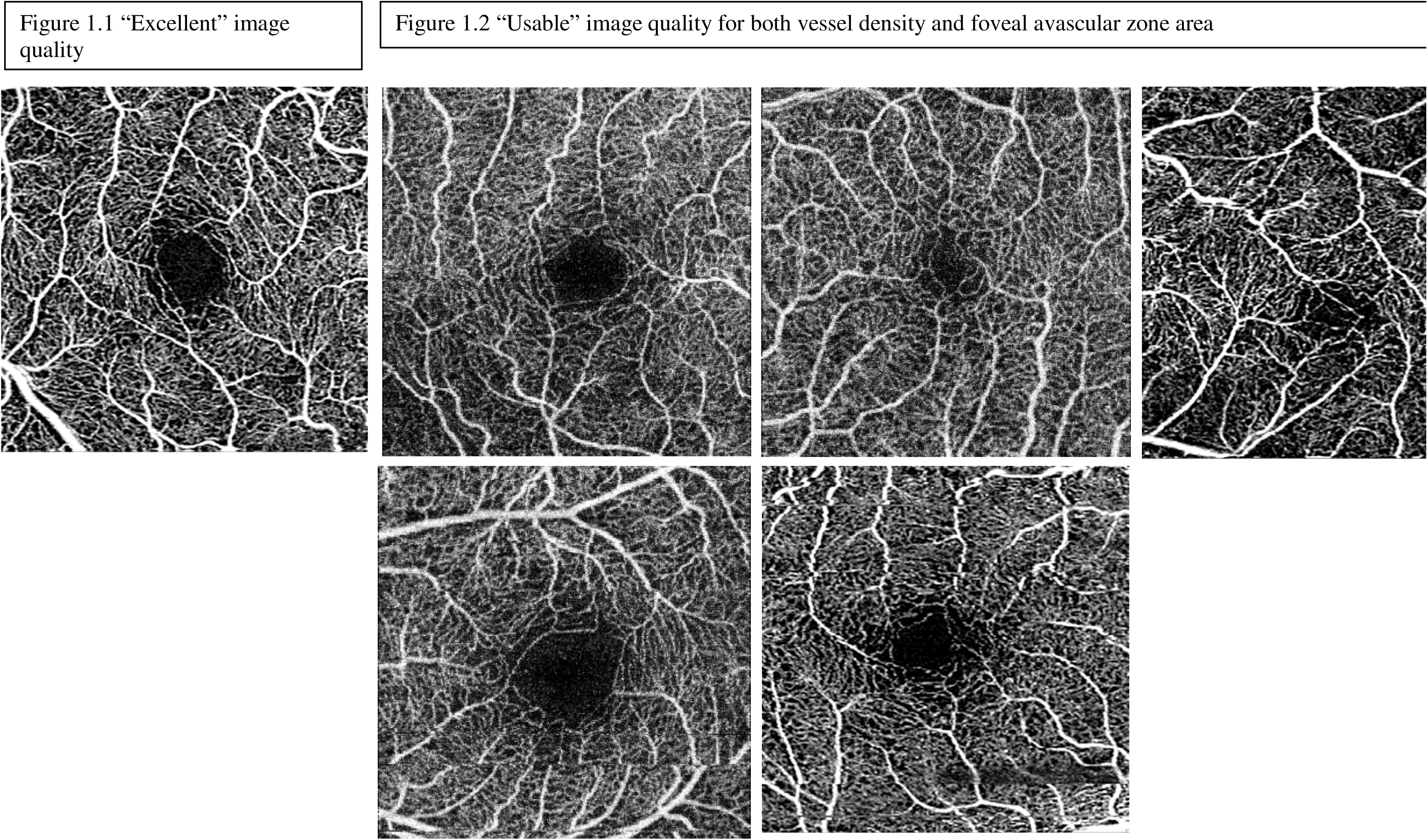
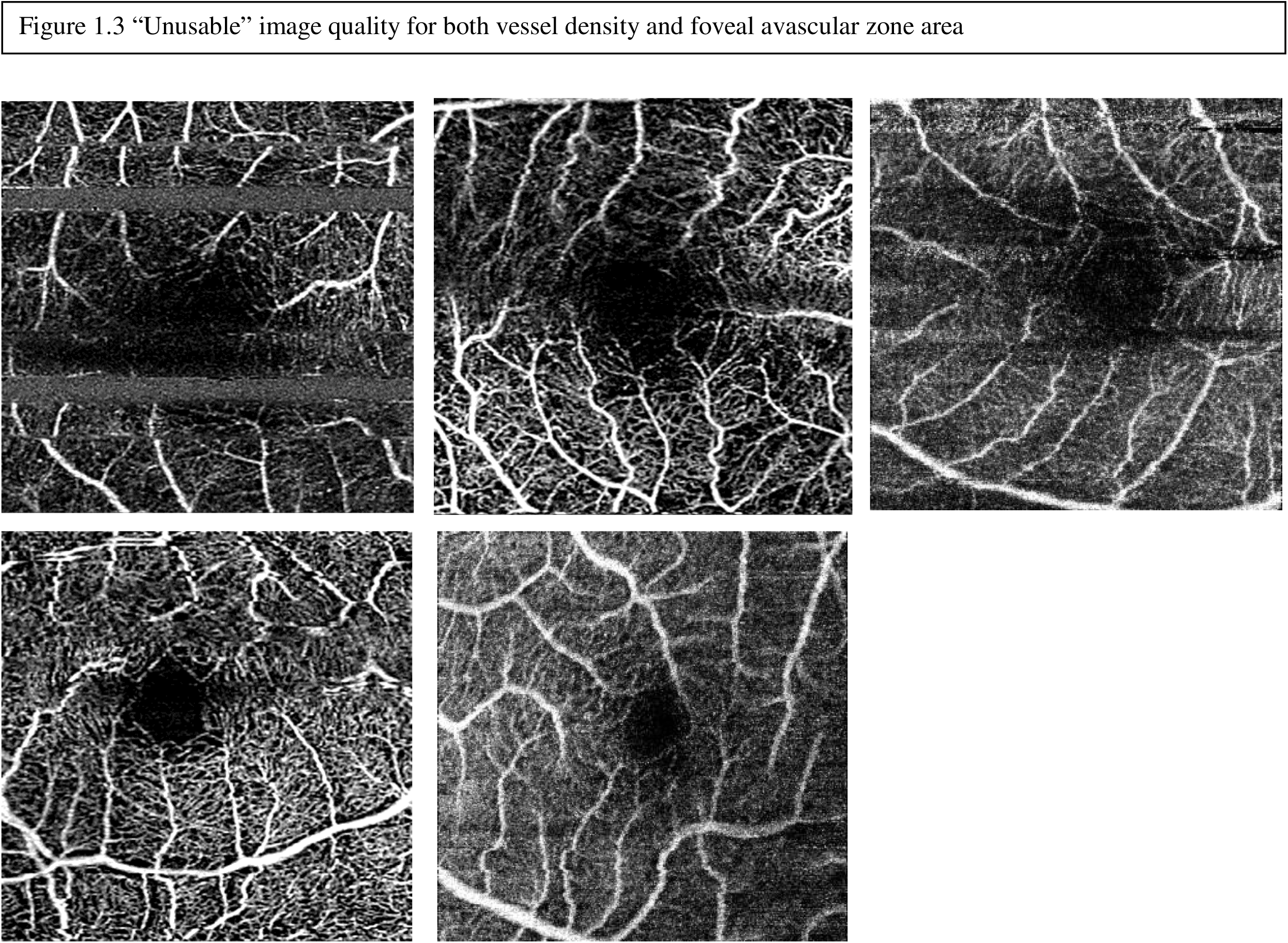
Exemplary images per category of OCT-A image quality (3×3-mm images)

#### Assessment of inter- and intragrader agreement for 3×3-mm OCT-A scans

Agreements for grading are shown in Table 2. All gradings are shown in Supplemental Material 2. For the intergrader agreement a total of 52 OCT-A images were assessed. The mean intergrader agreement for the overall image quality classified as “Unusable”, “Usable”, and “Excellent” image quality were 82.3%, 70.8%, and 87.1%, respectively. Concerning the vessel density and foveal avascular zone area, the mean intergrader agreements for “Unusable” were 80.0 % and 81.9%, respectively.

**Figure 2.**
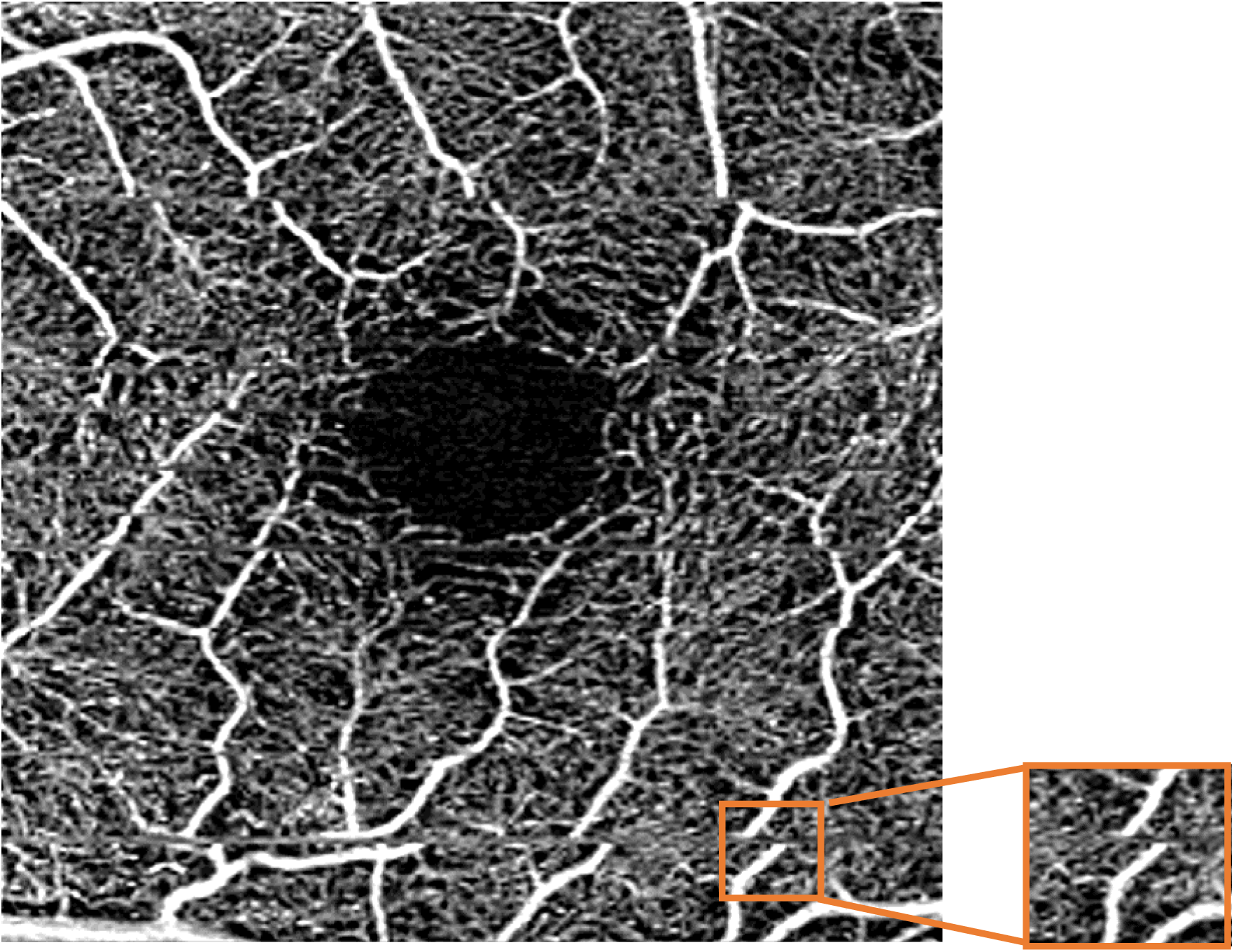
Exemplary 3×3-mm OCT-A image showing a vessel broken by full width (with z in). Other movement artefacts are also visible.

**Table 2.**
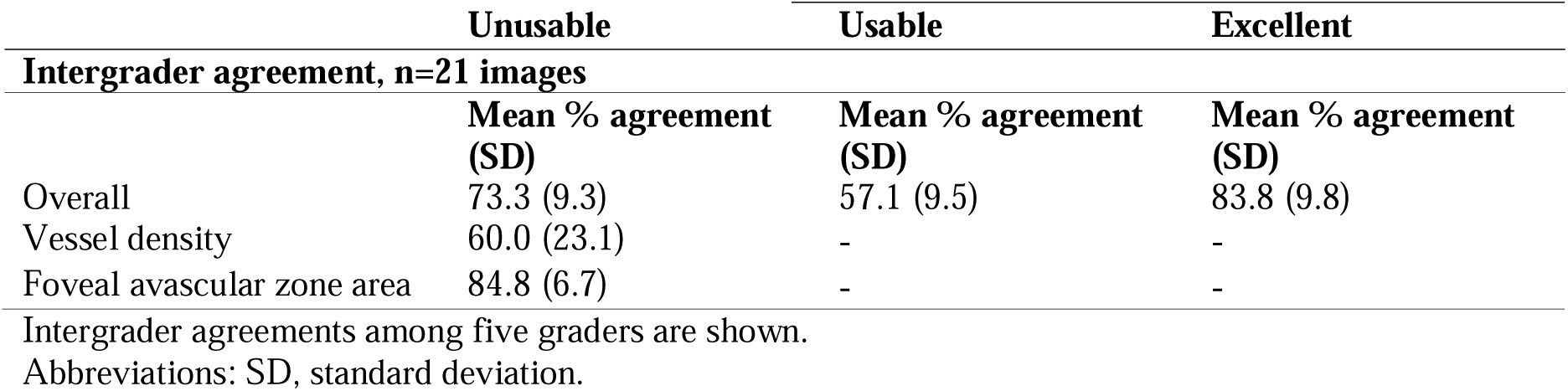
Inter- and intragrader agreement for “Unusable”, “Usable”, and “Excellent” image quality assessment of 3×3-mm OCT-A images.

For the intragrader agreement, a total of 27 OCT-A images were assessed twice by each grader. The mean intergrader agreements for overall “Unusable”, “Usable”, and “Excellent” image quality were 91.1%, 82.2%, and 81.9%, respectively. Concerning the vessel density and foveal avascular zone area, the mean intergrader agreements for “Unusable” were 89.6% and 92.6%, respectively.

#### Assessment of intergrader reliability for 6×6-mm OCT-A scans

A total of 21 OCT-A images were assessed. Agreements for grading are shown in Table 3. The mean intergrader agreement for the overall image quality <u>classified as</u> “Unusable”, “Usable”, and “Excellent” image quality were 73.3 %, 57.1%, and 83.8%, respectively. Concerning the vessel density and foveal avascular zone area, the mean intergrader agreements for “Unusable” were 60.0% and 84.8%, respectively.

**Table 3.**
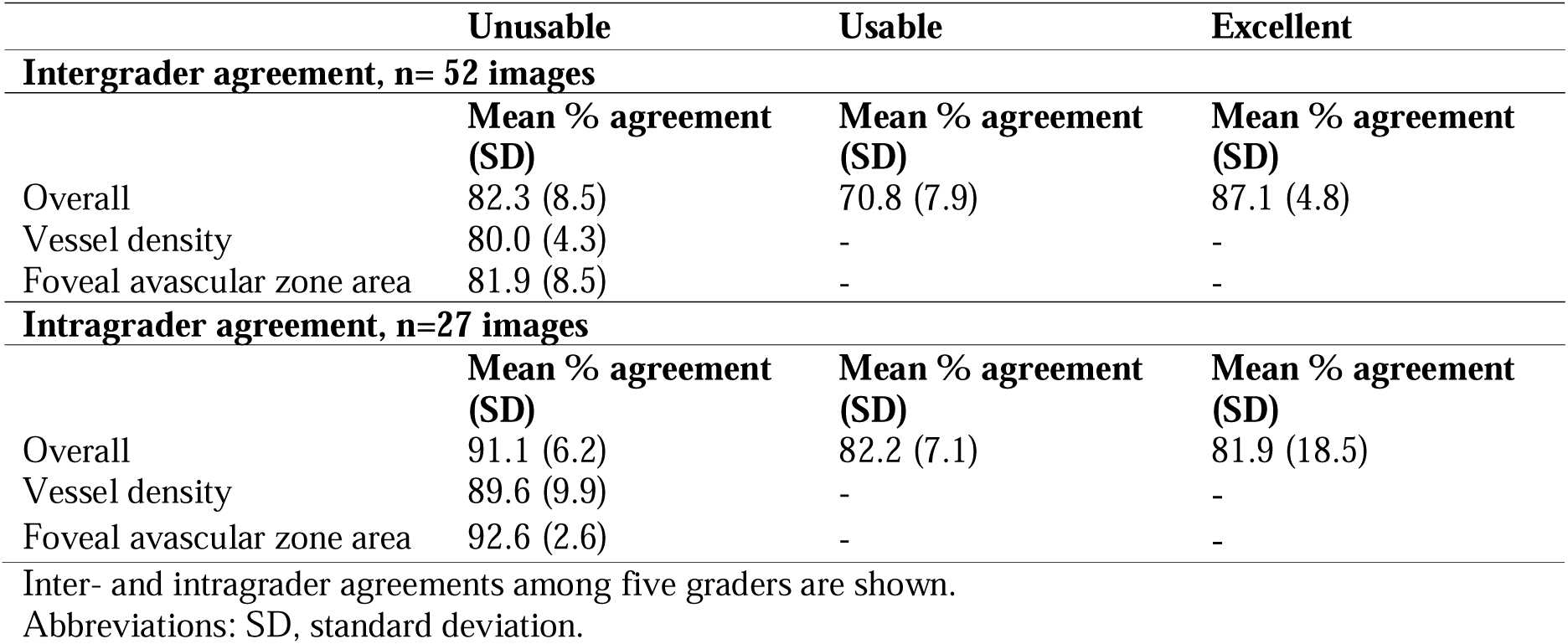
Intergrader agreement for “Unusable”, “Usable”, and “Excellent” image quality assessment of 6×6-mm OCT-A images.

#### Additional analyses

Results expressed as mean Cohen’s Kappa are shown in Tables S1 and S2. Intergrader agreements for overall “Unusable”, “Usable”, and “Excellent” image quality assessment for 3×3-mm OCT-A scans, expressed as mean Cohen’s kappa, were 0.64, 0.42, and 0.35, respectively (n=52 images). Intergrader agreements for “Unusable” vessel density and foveal avascular zone area of 3×3-mm OCT-A scans were 0.60 and 0.64, respectively. Intragrader agreements for overall “Unusable”, “Usable”, and “Excellent” image quality assessment for 3×3-mm OCT-A scans, were 0.77, 0.61, and 0.28, respectively (n=27 images). Intergrader agreements for “Unusable” vessel density and foveal avascular zone area of 3×3-mm OCT-A scans were 0.78 and 0.83, respectively. Intergrader agreements for overall “Unusable”, “Usable”, and “Excellent” image quality assessment for 6×6-mm OCT-A scans were 0.17, 0.06, 0.28, respectively (n=21 images). Intergrader agreements for “Unusable” vessel density and foveal avascular zone area of 6×6-mm OCT-A scans were 0.30 and 0.63, respectively.

### Part 2: Recommendations on epidemiological analyses using OCT-A imaging data

We developed a three-scenario framework that provides insight into selection and information bias due to suboptimal OCT-A image quality and ocular comorbidities (Panel 2). ^3–5^ We also provide recommendations for addressing potential selection bias.

<u>Scenario one: In the most common scenario</u> we suggest using a minimum set of OCT-A image quality criteria (i.e. least strict in terms of image quality assessment, in order to maximize the size of the study population for analyses whilst minimizing selection bias).

For this scenario, we propose to use the image quality grading criteria developed within this paper (i.e. excluding “Unusable” images, as defined in Panel 1). We note that using manufacturer-determined image quality indices such as “signal strength” or “signal-to-noise-ratio” thresholds are not a substitute for the grading criteria that we propose in this scenario or any other scenario.

We also recommend to use OCT-A images acquired in both eyes (if available), i.e. not restricting analyses to only the right or left eye. This will minimize the selection of participants and maximize statistical power.^10^ Researchers may consider to combine results of both eyes by averaging OCT-A results derived from the left and right eye, which may reduce the impact of measurement error. The biological grounding for this approach is that systemic risk factors that contribute to the pathobiology of many ocular diseases are biologically expected to have similar effects in both eyes.^9^ An alternative approach to using data from both eyes, without averaging results of both eyes, is multilevel analysis.^18^ Multilevel analysis allows to use data from both eyes, whilst efficiently accounting for the correlation between eyes within one individual. Although combining data on both eyes may be a suitable strategy for many research questions, we acknowledge that in certain specific situations (dependent on the research question) researchers may choose to consider to use data on one eye only. This may be the case when substantial differences are expected between eyes, e.g. if the microvasculature has been severely disrupted in one eye due to a disorder that does not affect both eyes equally and at the same time (this could be the case when studying retinal detachment for example).

<u>Scenario two:</u> In a second scenario, we propose to use more stringent image quality criteria (which reduces the chance of measurement error, i.e. information bias, but may increase the risk of selection bias).

For this scenario, we suggest four possible analytical strategies. We recommend use of these strategies after excluding “Unusable” images, as defined in Panel 1. First, we suggest to use only the best quality image from one eye for each subject. Second, we suggest to exclude all images with signal strength measures below the manufacturer recommended thresholds. Although such thresholds are likely arbitrary from one device to another, in general signal strength measures correlate well with image quality and are a good adjunct to the other quality control measures proposed here. Third, we suggest to adjust for signal strength by including signal strength as a covariate in the model.^19^ Adjustment for signal strength may improve precision of the estimate but may also lead to over adjustment, as signal strength is associated with OCT-A image quality, but not necessarily with the exposure or outcome variable.^19, 20^ Last, a fourth strategy is to exclude images of poor imaging quality as defined by the OSCAR-MP criteria.^11^ We recommend researchers use at least two of the proposed strategies.

Scenario three: In a third scenario, we suggest to evaluate the impact of ocular comorbidities on image quality measures (ocular comorbidities may negatively impact retinal capillary health and may predispose to poor quality OCT-A imaging).

For this scenario, we propose to exclude “Unusable” images (as defined in Panel 1) and in addition to conduct two analyses that provide insight into the impact of ocular comorbidities.

Ocular comorbidities that should, at minimum, be considered are age-related macular degeneration, glaucoma, retinopathy of any etiology, myopia, cataract, and corneal disease. We recommend conducting additional analyses in which 1) individuals with ocular comorbidities are excluded; and 2) ocular comorbidities are entered into the model as covariates. In our opinion it is important to conduct both analyses as they have different methodological underpinnings.^10^ Exclusion of individuals with ocular comorbidities may reduce measurement error but may induce selection bias.^10^ Adjustment for ocular comorbidities may reduce the chance of measurement error, but may (also) induce overadjustment bias (this may occur when ocular and systemic diseases with a shared pathobiology are entered in a model).^20^ We recommend researchers use both proposed strategies.

### Evaluation of scenarios and additional recommendations for addressing selection bias

A comparison of results from different scenarios will enable insight into the potential impact of information and selection bias due to poor OCT-A image quality. In the case results are similar across all scenarios the impact of information and selection bias due to OCT-A measurement quality may be minimal.

We recommend use of at least one of the following three strategies to address selection bias. A first approach is to evaluate characteristics of the individuals included and excluded in the different scenarios.^21^ If general characteristics of the populations differ, (some) selection bias may have occurred. A second approach is conducting analyses between non-missing variables (e.g. age or sex) and outcome(s) in the entire study population and in smaller populations (i.e. populations with sufficient OCT-A image quality). Comparison of results will provide insight into whether selection bias may have occurred.^21^ A third approach concerns use of inverse probability weighting, which allows to estimate associations whilst accounting for selectively missing data. This method uses weights (developed for prediction of non-missingness in the analytic sample) to account for selectively missing data. These weights are entered in the statistical model. More details are provided elsewhere.^22^

### Reporting

We recommend reporting results of all three analytical scenarios and addressing selection bias as proposed. For reasons of readability, certain scenarios are shown in the supplemental material.

### Panel 2 Three analytic scenarios

|  | Description |
| --- | --- |
| Scenario 1 | Minimal set of image quality criteria, to maximize study sample size |
| Scenario 2 | More stringent image quality than in scenario 1, recommended is to consider two of the following four options:<br>2.1) use only best quality image per individual<br>2.2) exclude images with signal strength based on manufacturer recommendations<br>2.3) adjustment for signal strength<br>2.4) exclude poor quality images based on OSCAR-MP criteria |
| Scenario 3 | Consider impact of ocular comorbidities on image quality, recommended is use both the following strategies:<br>3.1) exclusion of individuals with ocular comorbidities<br>3.2) adjustment for ocular comorbidities<br>Ocular comorbidities to consider are: age-related macular degeneration, glaucoma, retinopathy of any etiology, myopia, cataract, and corneal disease. |

## Discussion

In this manuscript researchers from the European Eye Epidemiology consortium and collaborators used an expert-based approach to develop a framework for assessment of OCT- A image quality and conducting analyses with OCT-A-derived variables in population-based studies. There are three main findings. One, the quality grading system developed using 3×3- mm OCT-A images showed substantial inter- and intragrader agreement for assessment of “Unusable” overall image quality, vessel density, and foveal avascular zone area. Two, the grading system showed substantial intergrader agreement for assessment of “Unusable” foveal avascular zone area on 6×6-mm scans, but lower agreement for assessment of overall quality and vessel density. Three, a methodological framework consisting of three scenarios to quantify impact of selection bias, information bias, and ocular comorbidities was developed.

This is the first study to provide a comprehensive framework for working with OCT-A images in population-based cohort studies. This work has an important added value to the already published OSCAR-MP criteria.^11^ One, we developed image quality criteria that are valid to use on devices of three of the most used OCT-A manufacturers (i.e. Heidelberg Engineering, Topcon, and Zeiss). In the previous study, only images acquired with a Heidelberg Engineering device were considered. Two, we developed criteria to determine whether images could be used for assessment of vessel density and foveal avascular zone area. Three, we evaluated grading criteria on 6×6-mm OCT-A images. Four, this study demonstrated validity of the grading system in the general population. The previous study only showed validity among patients with neurological diseases. Five, this study evaluated intragrader agreement for image quality assessment.

Intergrader agreement for the rejection of images with “Unusable” quality was similar when using our criteria (Cohen’s Kappa 0.64) and the OSCAR-MP criteria (Cohen’s Kappa 0.67). This is likely because both grading systems are in part similar.^11^ Both grading systems require the majority of “Usable” (or “Excellent”) images is free of artefacts. Our system is different from the OSCAR-MP on several points, 1) centration of the image is not considered in our criteria for assessment of image quality when the variable of interest is foveal avascular zone area; 2) the source of the error (i.e. type of artefact) is not specified in our system; and 3) no separate grading of retinal pathology is required. A potential advantage of our system is that it may be quicker to use (as less details are required to be documented).

Validation of the criteria of 6×6-mm images showed that our grading system, developed on 3×3-mm field-of-view images, performed relatively worse for the assessment of “Unusable” overall image quality; and for the assessment of “Unusable” vessel density on 6×6-mm images. A possible explanation is that the scan resolution for 6×6-mm images is generally lower than on 3×3-mm images, which makes it more difficult to assess capillary level detail (e.g. 6×6-mm images cover a larger retinal area than 3×3-mm images whilst fewer or the same number of B-scans are used to capture a 6×6-mm image [n=350 B-scans] than a 3×3-mm image [n=512 B-scans]).^1, 23^ This lower scan resolution is less likely to negatively impact the assessment of foveal avascular zone area size as the ‘absence’ of vasculature is used to assess foveal avascular zone area.^1, 23^ Indeed, intergrader agreement for usability of foveal avascular zone area was similar for 3×3-mm and 6×6-mm images (82% and 85%, respectively).

Although substantial percentage agreement was observed for “Excellent” quality images (82- 87%), relatively worse agreements were observed when Cohen’s kappa was calculated (0.28- 0.35). This may be due to an imbalanced distribution of different image quality gradings, as both rater agreement and prevalence are used in the calculation of Cohen’s Kappa. ^16, 17^ Indeed, ”Excellent” images are relatively less common than “Usable” or “Unusable” images. This phenomenon has previously been described; and is known as the “prevalence paradox”.^16^

Our findings have implications for future research. One, recommendations can be used to promote consistency in statistical analyses across population studies. Two, during the process of data collection exemplary images from this paper can be used to determine whether collected data are of sufficient quality. Three, our recommendations can be used for the development of artificial intelligence models for fully automatic assessment of OCT-A image quality.^24^ Four, given relatively lower intergrader agreement for assessment of vessel density in 6×6-mm images, developers of artificial intelligence algorithms may aim to develop automatic models for this.^24^ Open availability of a “benchmark” dataset containing images acquired using different protocols and on different devices would assist the development of such algorithms.

This study has certain strengths. One, OCT-A images collected in multiple countries and using different protocols and devices were used for the development of the image quality criteria. This implies that images of individuals from different ethnicities were considered (e.g. individuals of European and Indian descent).^10^ Also this implies that proposed image criteria can be used on images acquired from different devices with differences in resolution. Two, a relatively simple and practical grading system was developed, allowing for rapid assessment of OCT-A image quality. This is in particular relevant when large numbers of images are involved, such as in population studies. Three, we considered a three-level approach for assessment of overall image quality, allowing to distinguish between “excellent” and “usable” images. Advantages of this approach are that 1) a goal is provided to aim for during data acquisition; and 2) that in statistical analyses it is possible to only analyze those images free of measurement errors.

Limitations of this study are the following. One, we used an expert-based method to determine the percentage of the image that needs to be free of artefacts to be considered “Usable”. After intense debate, we settled on a subjective cut-off of 80% of the image required to be free of artefacts. Future studies may aim to provide quantitative insight into the impact of using differing thresholds of image quality free of artefacts. The quantitative impact on associations may be examined when e.g. studying risk factors for capillary deterioration; or associations with eye or systemic diseases. Two, we did not consider individuals of all ethnicities in this study (e.g. no individuals of African or Chinese descent were included). Whether findings are also valid in other populations requires further study.^10^ Three, we did not quantify intragrader agreement for the assessment of 6×6-mm OCT-A images.^10^ Four, there was some overlap in the 3×3-mm images used for development and validation of image quality criteria. Possibly some learning effect may have occurred, although the impact of any learning bias may have been minimized due to the presentation of images in a randomized order; and due to rotating or inverting of images graded twice. Five, we did not evaluate the impact of experience in grading OCT-A images or working with OCT-A images prior to grading image quality in this study.

In conclusion, in this study an expert-based approach was used to developed practical recommendations for quality assessment and use of OCT-A images in population-based studies. These recommendations provide a framework for future studies and aim to promote harmonization of analyses across studies.

## Acknowledgements

*Maastricht Study*

The authors would like to acknowledge ZIO foundation (Vereniging Regionale HuisartsenZorg Heuvelland) for their contribution to The Maastricht Study. The researchers are indebted to all participants for their willingness to participate in the study.

*Rotterdam Study*

The authors are grateful to the Rotterdam Study participants, the staff from the Rotterdam Study, and the participating general practitioners and pharmacists.

*Rhineland Study*

The authors would like to acknowledge the efforts of all study staff and participants in obtaining the OCTA data.

*Framingham Heart Study*

The authors would like to acknowledge the efforts of all study staff and participants in obtaining the OCTA data.

*SIGNATR*

The authors would like to acknowledge the SIGNATR Study Team: Gayatri Susarla, A Rizza, Ashley Li, Sam Han, Rehana Khan, Weilin Chan, Ines Lains, Atitaya Apivatthakakul, Kim Brustoski, Vikas Khetan, Robert Igo, Sudha K Iyengar, and Sinnakaruppan Mathavan. We would also like to acknowledge the participants who enrolled in the SIGNATR Study.

*Adolphe de Rothschild Foundation Hospital*

The authors would like to acknowledge Anne-Caroline Le Fur, Ozlem Erol, Justine Pineau and Justine Lafolie, from the French Image Reading Center, for their contributions to data collection.

## Author Contributions

F.C.T.v.d.H drafted the initial manuscript. F.C.T.v.d.H and A.H.K contributed to design, coordination, analyses, interpretation of the data, revised the manuscript critically for important intellectual content, and provided final approval of the version to be published. F.C.T.v.d.H also is the guarantor of this work and, as such, had full access to all the data in the study and takes responsibility for the integrity of the data and the accuracy of the data analyses. T.T.J.M.B., S.B. L.S., D.A.J., A.C.M., W.D.R., V.A.V., H.H. and A.H.K. contributed to development of image quality criteria, revised the manuscript critically for important intellectual content, and provided final approval of the version to be published. T.T.J.M.B. initiated the development of a joint statement. All other authors contributed to data collection, revised the manuscript critically for important intellectual content, and provided final approval of the version to be published

## Funding Sources

### The Maastricht Study

This study was supported by the European Regional Development Fund via OP-Zuid, the Province of Limburg, the Dutch Ministry of Economic Affairs (grant 31O.041), Stichting De Weijerhorst (Maastricht, the Netherlands), the Pearl String Initiative Diabetes (Amsterdam, the Netherlands), the Cardiovascular Center (CVC, Maastricht, the Netherlands), CARIM School for Cardiovascular Diseases (Maastricht, the Netherlands), CAPHRI School for Public Health and Primary Care (Maastricht, the Netherlands), NUTRIM School for Nutrition and Translational Research in Metabolism (Maastricht, the Netherlands), Stichting Annadal (Maastricht, the Netherlands), Health Foundation Limburg (Maastricht, the Netherlands), Perimed (Järfälla, Sweden), and by unrestricted grants from Janssen-Cilag B.V. (Tilburg, the Netherlands), Novo Nordisk Farma B.V. (Alphen aan den Rijn, the Netherlands), and Sanofi-Aventis Netherlands B.V. (Gouda, the Netherlands).

### Rotterdam Study

The Rotterdam Study is supported by the Algemene Nederlandse Vereniging ter Voorkoming van Blindheid, Oogfonds, Stichting voor Ooglijders, Stichting voor Blindenhulp, Henkes stichting, Rotterrdams Stichting voor Blindenbelangen, and Landelijke Stichting voor Blinden en Slechtzienden. Additional support was given by the Erasmus Medical Center, Erasmus University, Netherlands Organization for the Health Research and Development (ZonMw), the Research Institute for Diseases in the Elderly, the Ministry of Education, Culture and Science, the Ministry for Health, Welfare and Sports, the European Commission (DG XII), and the Municipality of Rotterdam.

### Rhineland Study

The Rhineland Study (P.I. Breteler) is primarily supported by DZNE core funding. The DZNE is funded by the Federal Ministry of Education and Research (BMBF) and the Ministry of Culture and Science of the German State of North Rhine-Westphalia.

### Framingham Heart Study

National Institutes of Health R01AG066524 (AHK, SS), FHS contract 75N92019D00031, and P30AG066546.

### SIGNATR

This work was supported by: National Eye Institute under R01 EY027134 and Government of India Department of Biotechnology under Grant BT/PR22701/MED/15/166/2016.

## Data Availability

Data are available for any researcher who meets the criteria for access to confidential data; the corresponding author may be contacted to request data.

## References

1. Kashani AH, Chen CL, Gahm JK, Zheng F, Richter GM, Rosenfeld PJ, Shi Y and Wang RK. Optical coherence tomography angiography: A comprehensive review of current methods and clinical applications. Prog Retin Eye Res. 2017;60:66–100.

2. Javed A, Khanna A, Palmer E, Wilde C, Zaman A, Orr G, Kumudhan D, Lakshmanan A and Panos GD. Optical coherence tomography angiography: a review of the current literature. J Int Med Res. 2023;51:3000605231187933.

3. Werner AC and Shen LQ. A Review of OCT Angiography in Glaucoma. Semin Ophthalmol. 2019;34:279–286.

4. Courtie E, Kirkpatrick JRM, Taylor M, Faes L, Liu X, Logan A, Veenith T, Denniston AK and Blanch RJ. Optical coherence tomography angiography analysis methods: a systematic review and meta-analysis. Sci Rep. 2024;14:9643.

5. Spaide RF, Fujimoto JG, Waheed NK, Sadda SR and Staurenghi G. Optical coherence tomography angiography. Prog Retin Eye Res. 2018;64:1–55.

6. Kashani AH, Asanad S, Chan JW, Singer MB, Zhang J, Sharifi M, Khansari MM, Abdolahi F, Shi Y, Biffi A, Chui H and Ringman JM. Past, present and future role of retinal imaging in neurodegenerative disease. Prog Retin Eye Res. 2021;83:100938.

7. Ibrahim Y, Xie J, Macerollo A, Sardone R, Shen Y, Romano V and Zheng Y. A Systematic Review on Retinal Biomarkers to Diagnose Dementia from OCT/OCTA Images. J Alzheimers Dis Rep. 2023;7:1201–1235.

8. Zhang JF, Wiseman S, Valdes-Hernandez MC, Doubal FN, Dhillon B, Wu YC and Wardlaw JM. The Application of Optical Coherence Tomography Angiography in Cerebral Small Vessel Disease, Ischemic Stroke, and Dementia: A Systematic Review. Front Neurol. 2020;11:1009.

9. Kellner RL, Harris A, Ciulla L, Guidoboni G, Verticchio Vercellin A, Oddone F, Carnevale C, Zaid M, Antman G, Kuvin JT and Siesky B. The Eye as the Window to the Heart: Optical Coherence Tomography Angiography Biomarkers as Indicators of Cardiovascular Disease. J Clin Med. 2024;13.

10. Ahlbom A. Modern Epidemiology, 4th edition. TL Lash, TJ VanderWeele, S Haneuse, KJ Rothman. Wolters Kluwer, 2021. Eur J Epidemiol. 2021;36:767–768.

11. Wicklein R, Yam C, Noll C, Aly L, Banze N, Romahn EF, Wolf E, Hemmer B, Oertel FC, Zimmermann H, Albrecht P, Ringelstein M, Baumann C, Feucht N, Penkava J, Havla J, Gernert JA, Mardin C, Vasileiou ES, Van Der Walt A, Al-Louzi O, Cabello S, Vidal-Jordana A, Kramer J, Wiendl H, Preiningerova JL, Ciccarelli O, Garcia-Martin E, Kana V, Calabresi PA, Paul F, Saidha S, Petzold A, Toosy AT, Knier B and Consortium I. The OSCAR-MP Consensus Criteria for Quality Assessment of Retinal Optical Coherence Tomography Angiography. Neurol Neuroimmunol Neuroinflamm. 2023;10.

12. Schram MT, Sep SJ, van der Kallen CJ, Dagnelie PC, Koster A, Schaper N, Henry RM and Stehouwer CD. The Maastricht Study: an extensive phenotyping study on determinants of type 2 diabetes, its complications and its comorbidities. Eur J Epidemiol. 2014;29:439–51.

13. Ikram MA, Brusselle G, Ghanbari M, Goedegebure A, Ikram MK, Kavousi M, Kieboom BCT, Klaver CCW, de Knegt RJ, Luik AI, Nijsten TEC, Peeters RP, van Rooij FJA, Stricker BH, Uitterlinden AG, Vernooij MW and Voortman T. Objectives, design and main findings until 2020 from the Rotterdam Study. Eur J Epidemiol. 2020;35:483–517.

14. Tsao CW and Vasan RS. Cohort Profile: The Framingham Heart Study (FHS): overview of milestones in cardiovascular epidemiology. Int J Epidemiol. 2015;44:1800–13.

15. Susarla G, Rizza AN, Li A, Han S, Khan R, Chan W, Lains I, Apivatthakakul A, Brustoski K, Khetan V, Raman R, Igo RP, Jr., Iyengar SK, Mathavan S and Sobrin L. Younger Age and Albuminuria are Associated with Proliferative Diabetic Retinopathy and Diabetic Macular Edema in the South Indian GeNetics of DiAbeTic Retinopathy (SIGNATR) Study. Curr Eye Res. 2022;47:1389–1396.

16. Byrt T, Bishop J and Carlin JB. Bias, prevalence and kappa. J Clin Epidemiol. 1993;46:423–9.

17. Delgado R and Tibau XA. Why Cohen’s Kappa should be avoided as performance measure in classification. PLoS One. 2019;14:e0222916.

18. Hofoss D, Veenstra M and Krogstad U. Multilevel analysis in health services research: a tutorial. Ann Ist Super Sanita. 2003;39:213–22.

19. Yu JJ, Camino A, Liu L, Zhang X, Wang J, Gao SS, Jia Y and Huang D. Signal Strength Reduction Effects in OCT Angiography. Ophthalmol Retina. 2019;3:835–842.

20. Schisterman EF, Cole SR and Platt RW. Overadjustment bias and unnecessary adjustment in epidemiologic studies. Epidemiology. 2009;20:488–95.

21. Hernan MA, Hernandez-Diaz S and Robins JM. A structural approach to selection bias. Epidemiology. 2004;15:615–25.

22. Chesnaye NC, Stel VS, Tripepi G, Dekker FW, Fu EL, Zoccali C and Jager KJ. An introduction to inverse probability of treatment weighting in observational research. Clin Kidney J. 2022;15:14–20.

23. Ho J, Dans K, You Q, Nudleman ED and Freeman WR. Comparison of 3mm x 3mm versus 6mm x 6mm optical coherence tomography angiography scan sizes in the evaluation of non-proliferative diabetic retinopathy. Retina. 2019;39:259–264.

24. Ting DSW, Pasquale LR, Peng L, Campbell JP, Lee AY, Raman R, Tan GSW, Schmetterer L, Keane PA and Wong TY. Artificial intelligence and deep learning in ophthalmology. Br J Ophthalmol. 2019;103:167–175.

