## Supplemental Material 1 for "Retinal optical coherence tomography angiography imaging in population studies"

**Table of Contents**

**Supplemental Methods**

**Supplemental Figures**

-Figure S1 Exemplary images of 3x3-mm OCT-A scans

-Figure S2 Exemplary images of 6x6-mm OCT-A scans
-Figure S3 Exemplary 3x3-mm OCT-A scans for excellent, usable, and unusable overall image quality per device type
*-*Figure S4 Exemplary 6x6-mm OCT-A scans for excellent, usable, and unusable overall image quality acquired with the CIRRUS HD-OCT 5000 (Zeiss Meditec. Inc., Dublin, CA, USA)

**Supplemental Tables**
*-*Table S1 Inter- and intragrader agreement for “Unusable”, “Usable”, and “Excellent” image quality assessment of 3x3-mm OCT-A images, expressed using Cohen’s kappa
-Table S2 Intergrader agreement for “Unusable”, “Usable”, and “Excellent” image quality assessment of 6x6-mm OCT-A images

**Supplemental Methods**

### 1. Maastricht Study

The Maastricht Study is an observational, population-based cohort study that consists of 9,187 participants.^19^ Eligible for participation were all individuals aged between 40 and 75 years old and living in the southern part of the Netherlands. Recruitment was stratified according to known type 2 diabetes status, with an oversampling of individuals with type 2 diabetes, for reasons of efficiency. Data collection started in November 2010. OCT-A imaging was added as part of repeat measurements, starting from January 2020. All participants gave written consent for participation.

### 2. Rotterdam Study

The Rotterdam Study (RS) is a prospective population-based cohort study of residents living in Ommoord, a district of the city of Rotterdam, The Netherlands^1^. The RS consists of four cohorts. The first cohort (RS-I) started in 1991 and consisted of 7,983 participants >55 years old (range 55.0 – 99.2, response rate of 78%). The second cohort (RS-II) started recruiting in 2000 and consisted of 3,011 participants >55 years old (range 55.2 – 98.9, response rate of 67.3%). The third cohort (RS-III) included people aged >45 years old (range 45.7 - 90.1) and consisted of 3,932 participants (response rate 64.9%) starting from the year 2006. The fourth cohort (RS-IV) started in 2016, included people aged 40 years and over (range 49.0 – 63.0), and consisted of 3,005 participants (response rate 45.4%). Collection of OCT-A images started from December 2021. All participants gave written informed consent for participation.

### 3. Rhineland Study

The Rhineland Study is an ongoing population-based cohort study that aims to recruit up to 20,000 participants. Eligible for participation are individuals aged 30 years or over and living in either of 2 geographically defined areas in Bonn, Germany. Data collection started in 2016. Until July 2024, more than 11,000 individuals participated. Collection of OCT-A images started from March 2023. A pilot study in the Rhineland Study has been conducted to collect 5 participants’ bilateral OCT-A images for this particular study. All participants gave written consent for participation.

### 4. Framingham Heart Study

The Framingham Heart Study is an observational, population-based cohort study that consists of 1,415 eligible participants who attended Exam 10 and 5. Eligible for participation were all individuals aged between 30 and 62 years old and living in Framingham, Massachusetts, United States. Data collection started in 1948. Collection of OCT-A images started from 2020. All participants gave written consent for participation.

### 5. SIGNATR Study

The South Indian GeNetics of DiAbeTic Retinopathy (SIGNATR) Study is an observational, cohort study that presently consists of n=2,941 participants. Eligible for participation were all individuals with type 2 diabetes living in one of the four states of South India. Data collection started in 2018. OCT-A images were collected among n=348 individuals. All participants gave written consent for participation.

**Supplemental Figures**

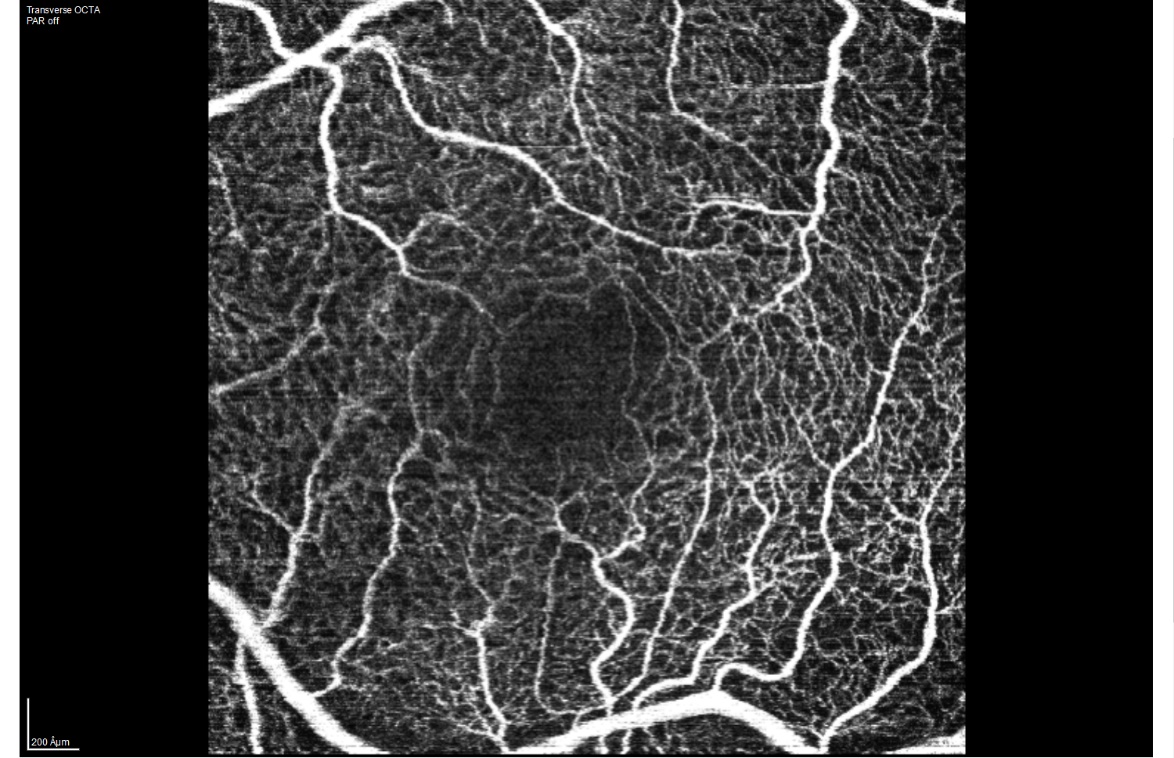

*Figure S1.1 3x3-mm fovea-centered OCT-A image in the Maastricht Study, acquired with the Heidelberg Spectralis (Heidelberg Engineering, Heidelberg, Germany).*

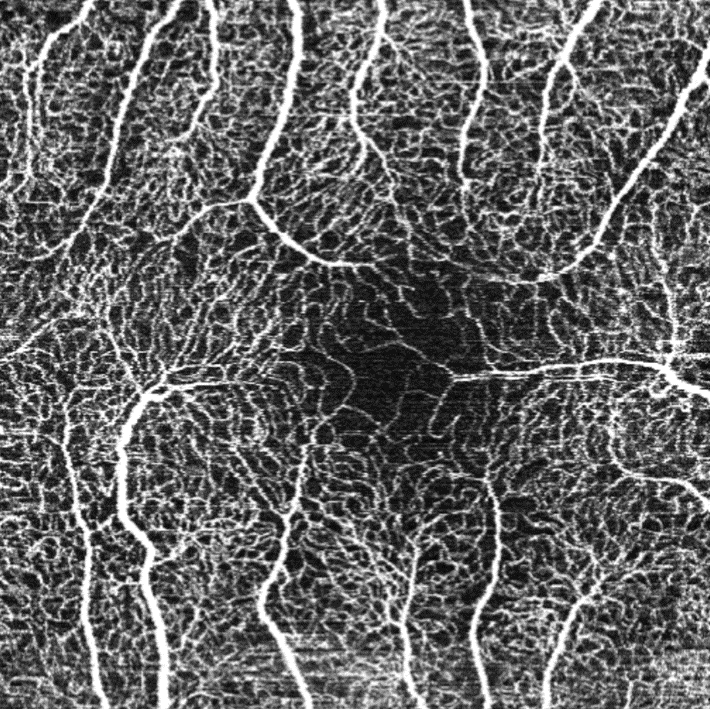

*Figure S1.2 3x3-mm fovea-centered OCT-A image in the Rhineland Study, acquired with the Heidelberg Spectralis (Heidelberg Engineering,* *Heidelberg, Germany).*

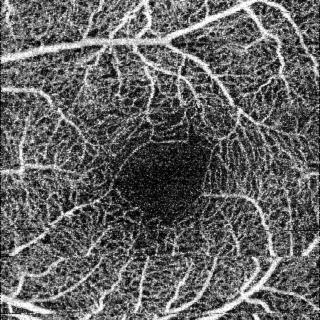

*Figure S1.3 3x3-mm fovea-centered OCT-A image in the Rotterdam Study, acquired with Triton (Topcon, Tokyo, Japan).*

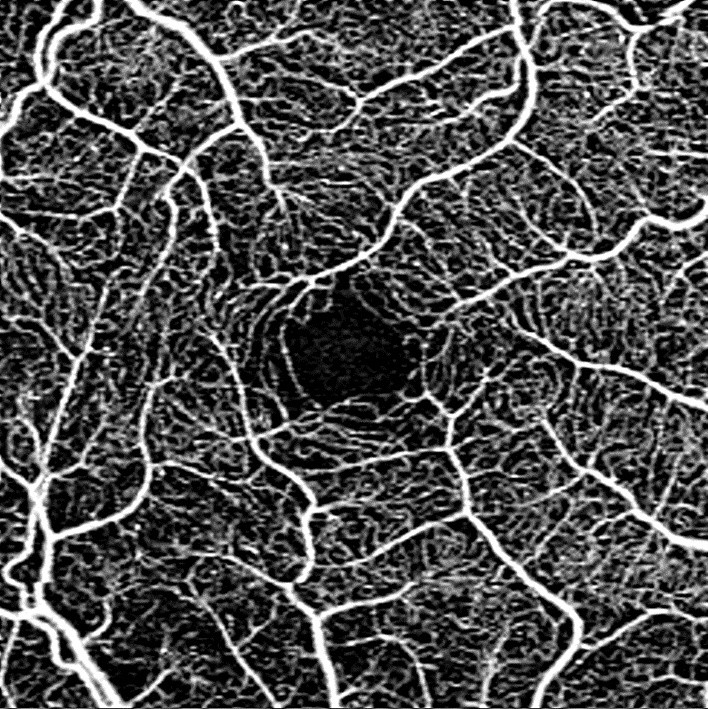

*Figure S1.4 3x3-mm fovea-centered OCT-A image in the Framingham Heart Study, acquired acquired with the CIRRUS 6000 (Angioplex, Zeiss Meditec. Inc., Dublin, CA, USA).*

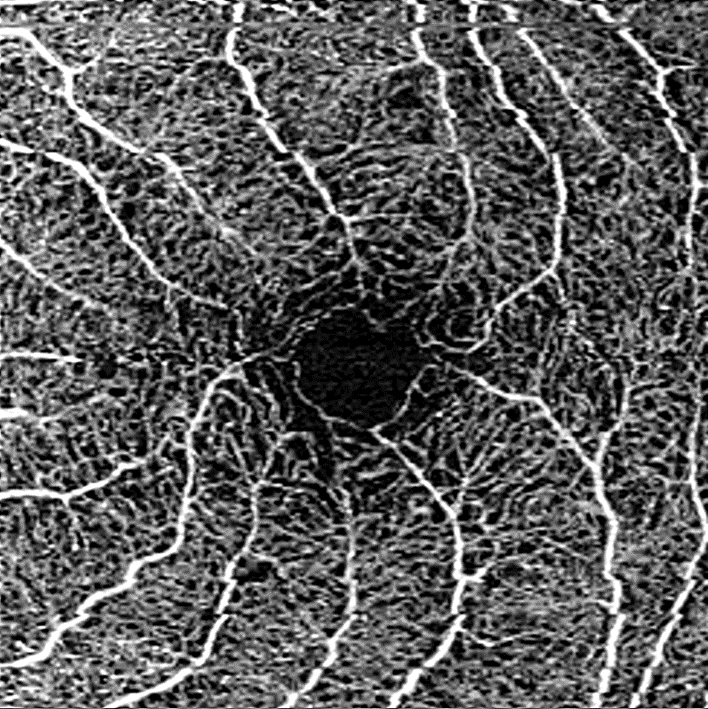

*Figure S1.5 3x3-mm fovea-centered OCT-A image from Adolphe de Rothschild Foundation Hospital, acquired with the CIRRUS Model 6000 (Angioplex, Zeiss Meditec. Inc., Dublin, CA, USA).*

**Figure S1 Exemplary images of 3x3-mm OCT-A scans**

Abbreviations: OCT-A, optical coherence tomography angiography.

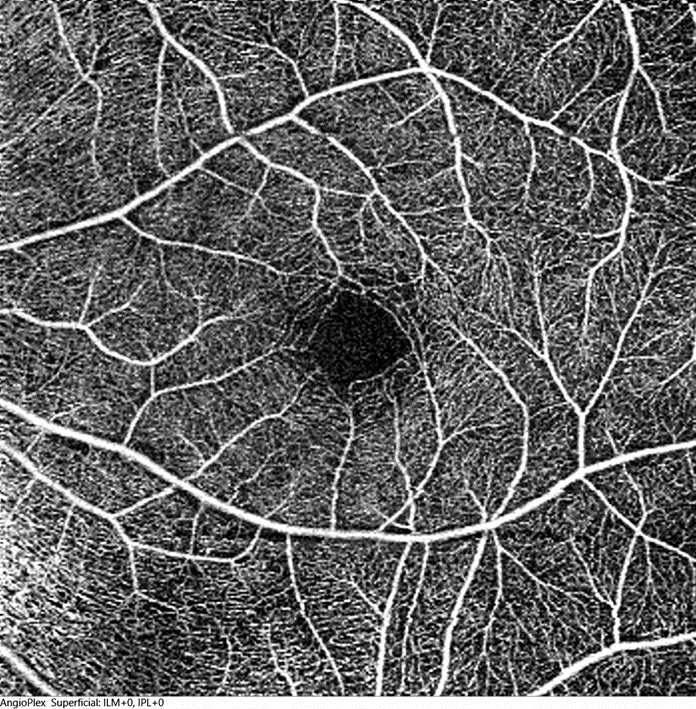

*Figure S2.1 6x6-mm fovea-centered OCT-A image in the SIGNATR Study, acquired with CIRRUS HD-OCT 5000 (Zeiss Meditec. Inc., Dublin, CA, USA).*

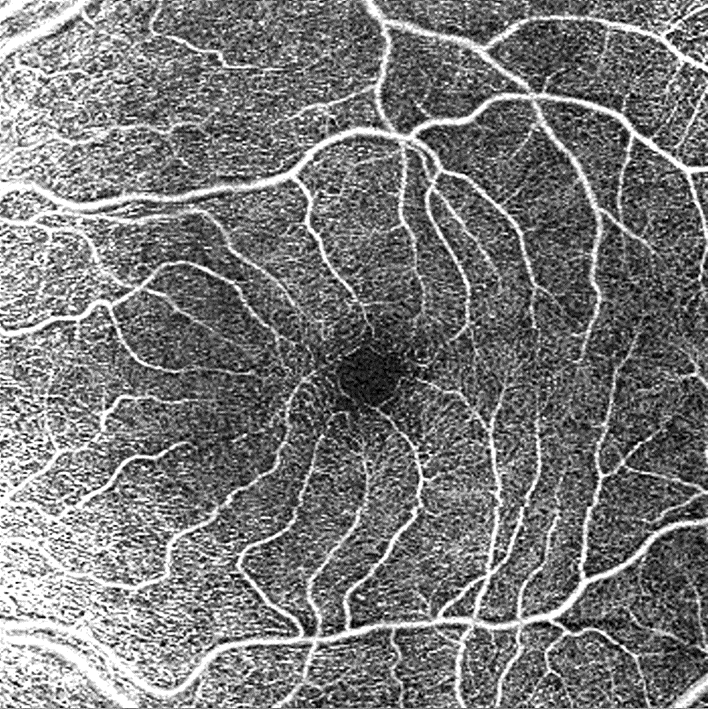

*Figure S2.2 6x6-mm fovea-centered OCT-A image from Adolphe de Rothschild Foundation Hospital, acquired with the CIRRUS Model 6000 (Angioplex, Zeiss, Meditec. Inc., Dublin, CA, USA).*

**Figure S2 Exemplary images of 6x6-mm OCT-A scans**

Abbreviations: OCT-A, optical coherence tomography angiography.

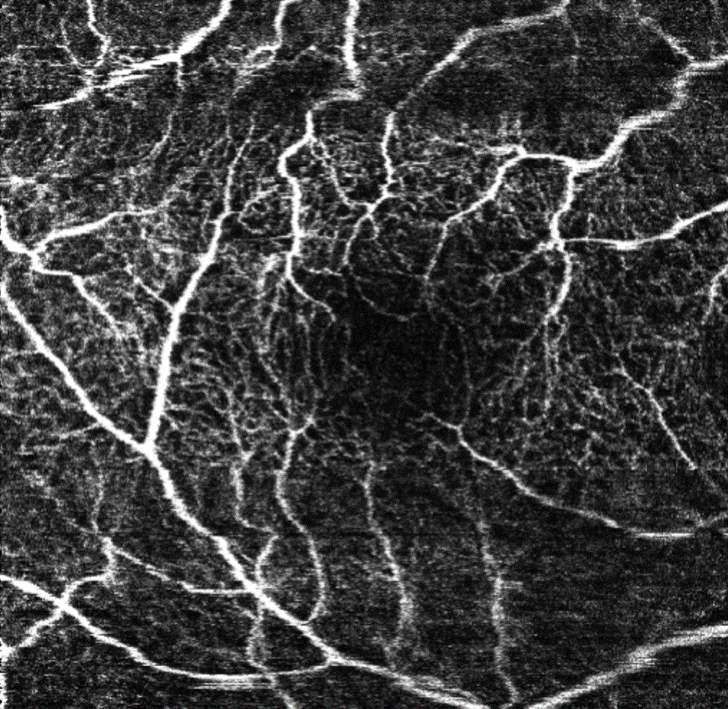

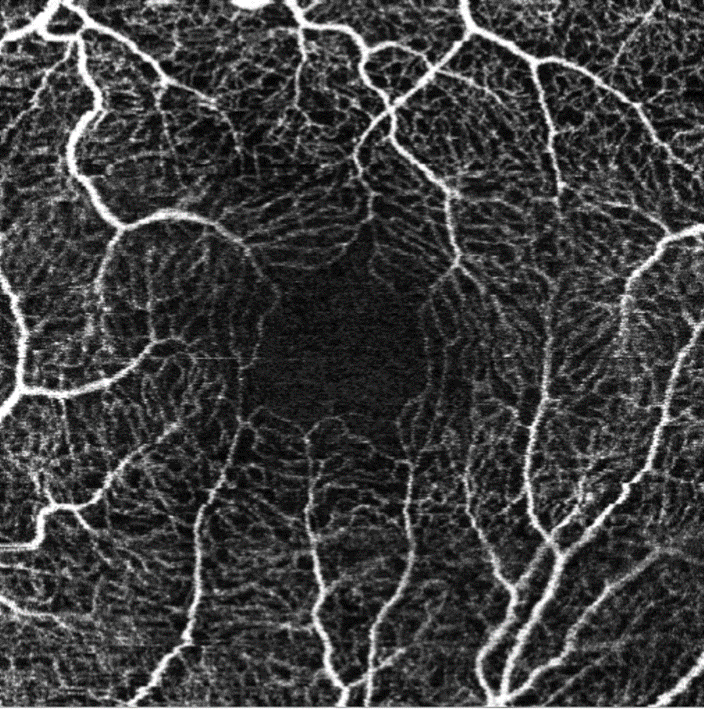

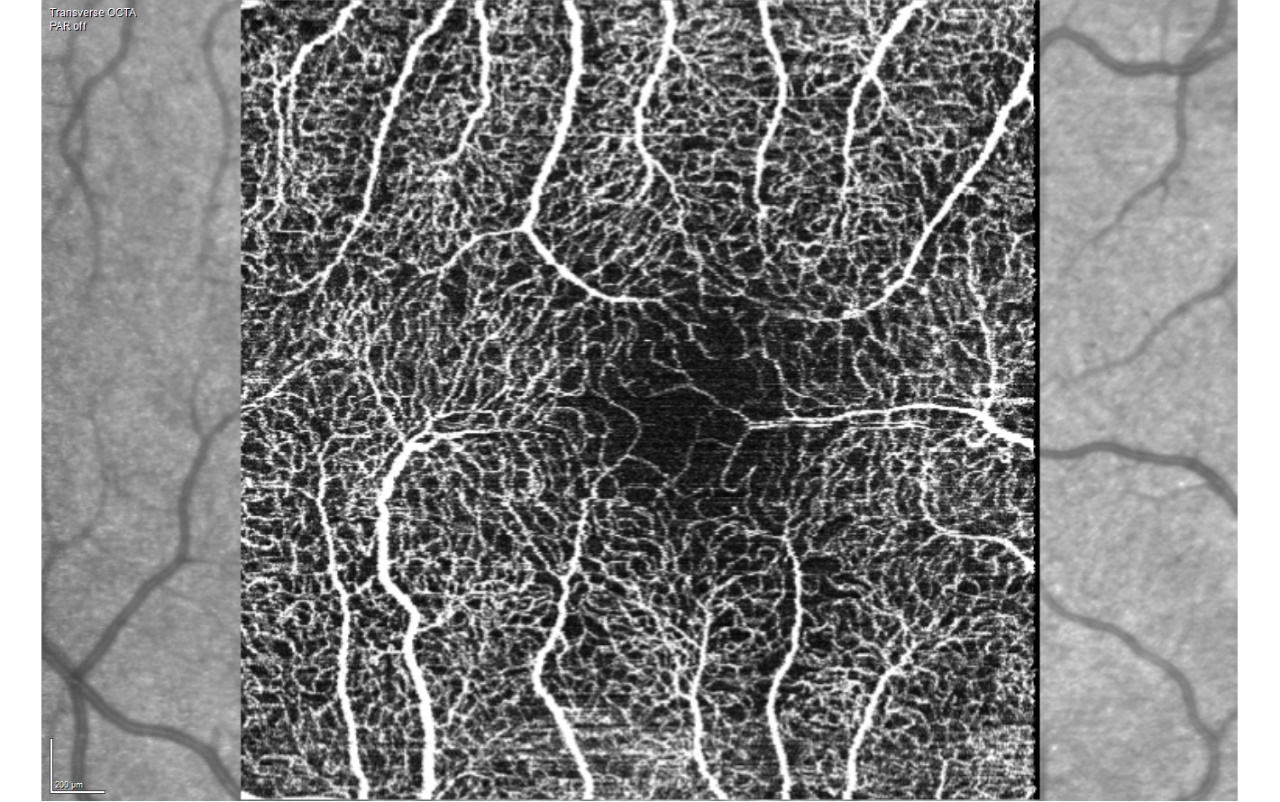

Unusable

Usable

Excellent

*Figure S3.1 examples of 3x3-mm fovea-centered OCT-A image acquired with the Heidelberg Spectralis (Heidelberg Engineering, Heidelberg, Germany).*

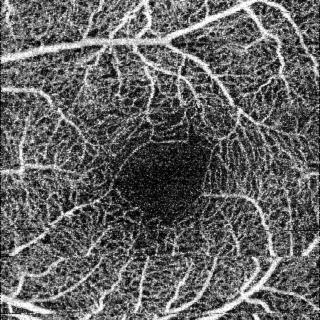

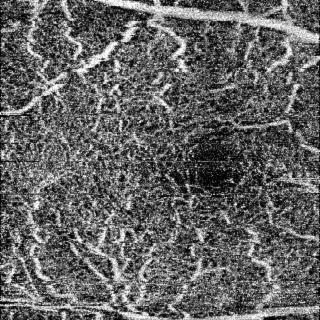

Unusable

Usable

*Figure S3.2 examples of 3x3-mm fovea-centered OCT-A images acquired with Triton (Topcon, Tokyo, Japan).*

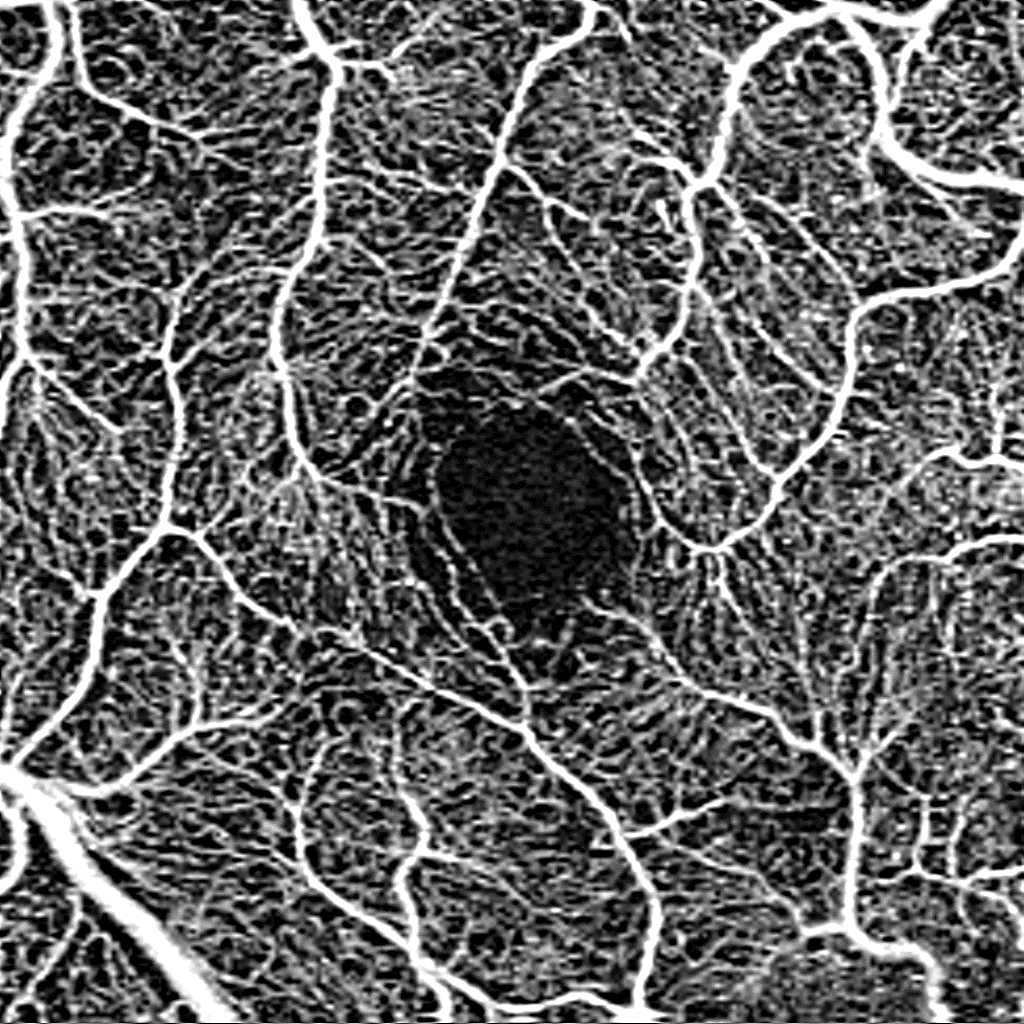

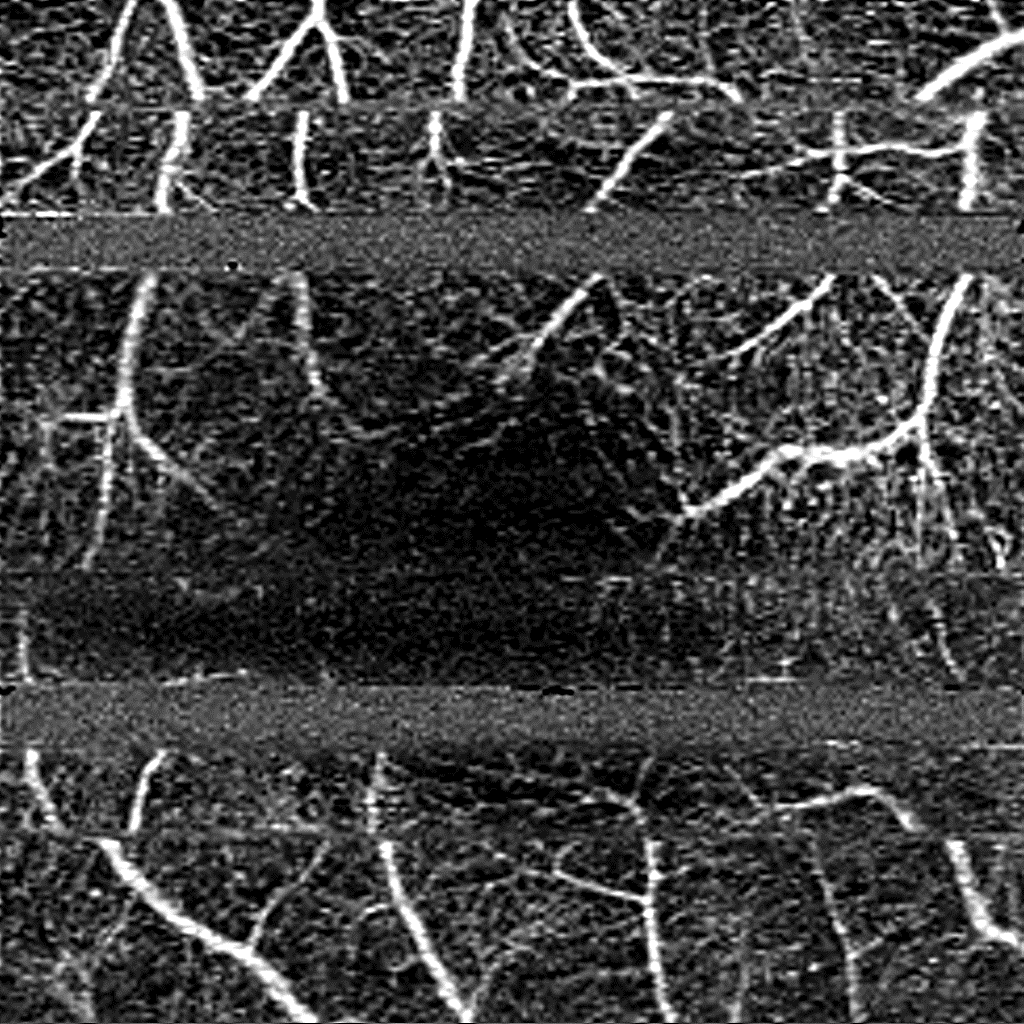

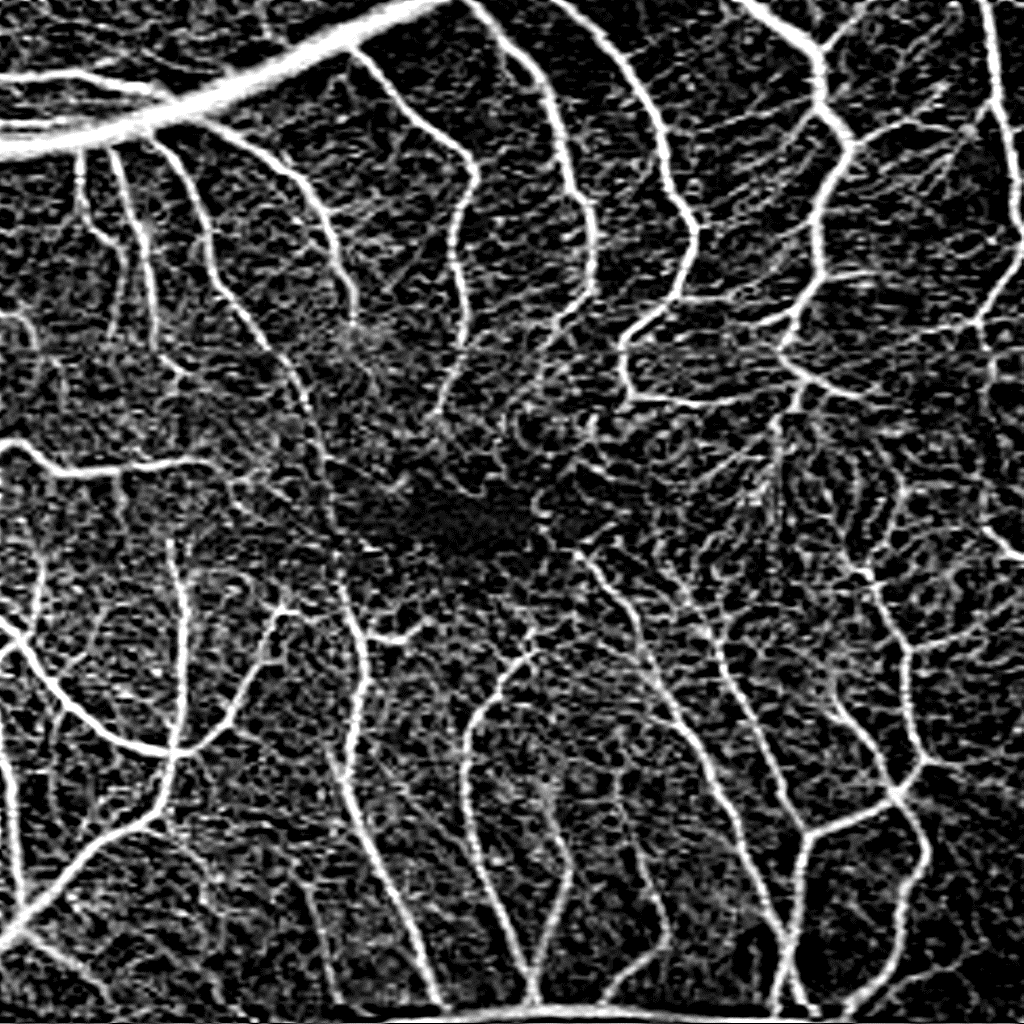

Excellent

Usable

Unusable

*Figure S3.3 3x3-mm fovea-centered OCT-A images, acquired acquired with CIRRUS Model 6000 (Angioplex, Zeiss Meditec. Inc., Dublin, CA, USA).*

**Figure S3 Exemplary 3x3-mm OCT-A scans for excellent, usable, and unusable overall image quality per device type**

Abbreviations: OCT-A, optical coherence tomography angiography. For Topcon images no excellent images were available.

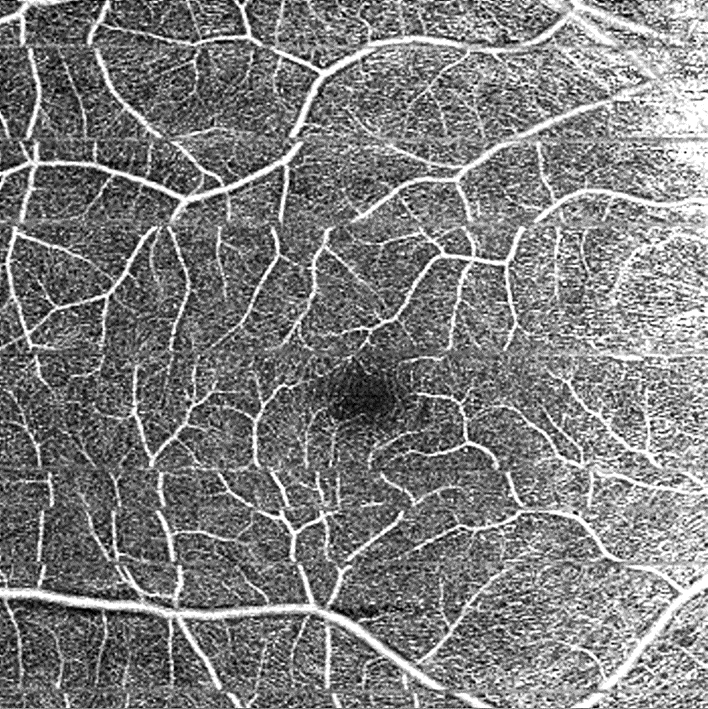

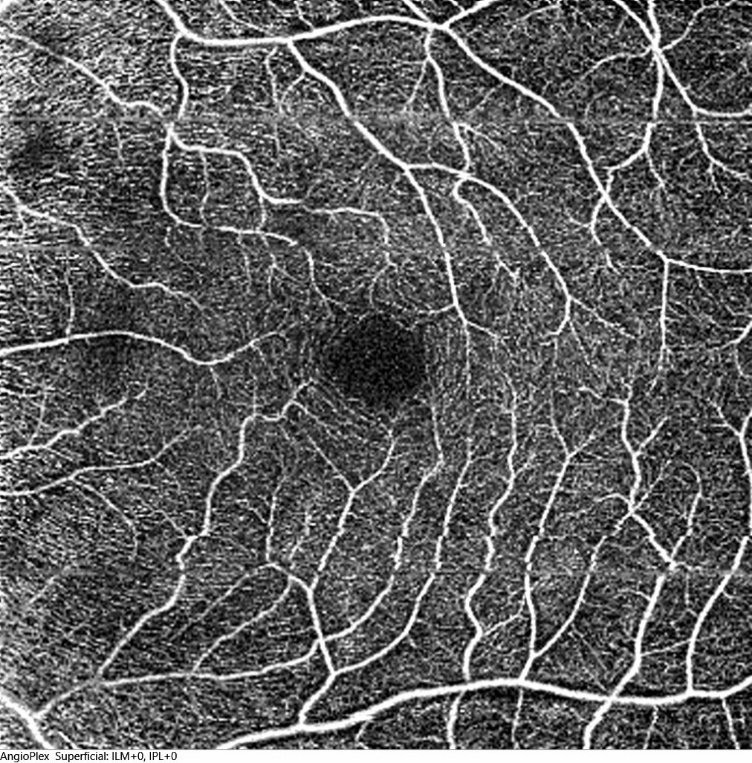

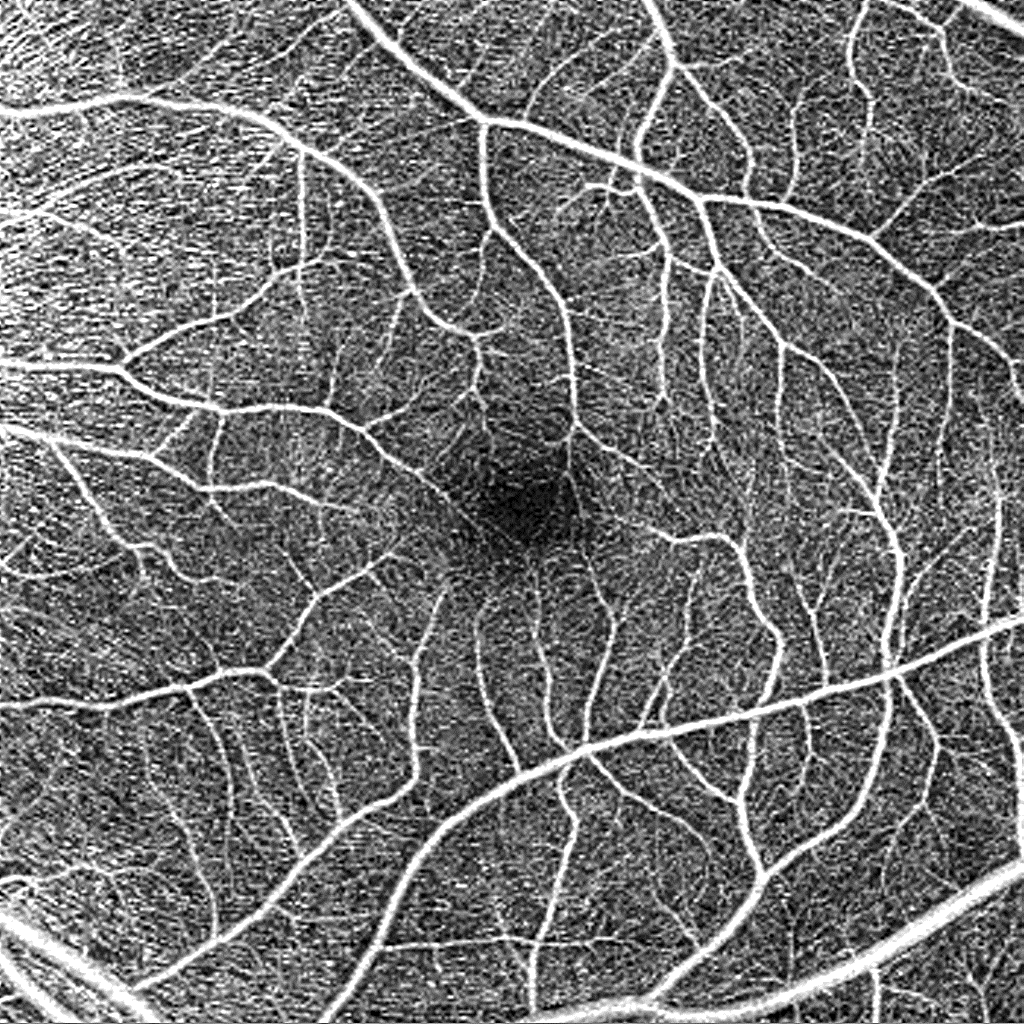

Usable

Excellent

Unusable

**Figure S4 Exemplary 6x6-mm OCT-A scans for excellent, usable, and unusable overall image quality acquired with the CIRRUS Model 5000 or 6000 (HD-OCT or Angioplex, Zeiss Meditec. Inc., Dublin, CA, USA)**

Abbreviations: OCT-A, optical coherence tomography angiography.

**Table S1 Inter- and intragrader agreement for “Unusable”, “Usable”, and “Excellent” image quality assessment of 3x3-mm OCT-A images, expressed using Cohen’s kappa**

|  | **Unusable** | **Usable** | **Excellent** |
| --- | --- | --- | --- |
| **Intergrader agreement, n= 52 images** | | | |
|  | **Mean Cohen’s kappa (SD)** | **Mean Cohen’s kappa (SD)** | **Mean Cohen’s kappa (SD)** |
| Overall | 0.64 (0.15) | 0.42 (0.16) | 0.35 (0.29) |
| Vessel density | 0.60 (0.08) | **-** | **-** |
| Foveal avascular zone area | 0.64 (0.15) | **-** | **-** |
| **Intragrader agreement, n=27 images** | | | |
|  | **Mean % agreement (SD)** | **Mean % agreement (SD)** | **Mean % agreement (SD)** |
| Overall | 0.77 (0.20) | 0.61 (0.21) | 0.28 (0.38) |
| Vessel density | 0.78 (0.20) | **-** | **-** |
| Foveal avascular zone area | 0.83 (0.05) | **-** | **-** |

Inter- and intragrader agreements among five graders are shown.
Abbreviations: SD, standard deviation.

**Table S2 Intergrader agreement for “Unusable”, “Usable”, and “Excellent” image quality assessment of 6x6-mm OCT-A images**

|  | **Unusable** | **Usable** | **Excellent** |
| --- | --- | --- | --- |
| **Intergrader agreement, n=21 images** | | | |
|  | **Mean % agreement (SD)** | **Mean % agreement (SD)** | **Mean % agreement (SD)** |
| Overall | 0.17 (0.16) | 0.06 (0.15) | 0.28 (0.28) |
| Vessel density | 0.30 (0.26) | **-** | **-** |
| Foveal avascular zone area | 0.63 (0.13) | **-** | **-** |

Intergrader agreements among five graders are shown.
Abbreviations: SD, standard deviation.
